# Questioning the Benchmarks: Hidden Circularity in AQ-10 Autism Screening Datasets

**DOI:** 10.64898/2026.11.01.26367003

**Authors:** Soham Biren Katlariwala, Amina Saeed, Harina Kumari Khati, Nirban Roy, Mahdisa Moradi

**Affiliations:** Dept. of Computer Engineering, Gujarat Technological University, Ahmedabad, Gujarat 382424, India; Dept. of Health Science, University of the People, Pasadena, CA 91101, USA; Dept. of CS & IT, Virtual University of Pakistan, Lahore 54000, Pakistan; Dept. of Computer Science and Information Technology, Padma Kanya Multiple Campus, Tribhuvan University, Kathmandu 44600, Nepal; Dept. of Computer Science & Engineering, Institute of Engineering & Management, Kolkata, India

**Keywords:** autism spectrum disorder, label leakage, dataset circularity, tabular deep learning, screening instruments, AQ-10, DASS-42, psychometric data quality

## Abstract

Autism spectrum disorder (ASD) screening increasingly relies on machine learning models trained on the AQ-10 and Q-CHAT questionnaire datasets, and these models are widely reported to reach 95 to 99 percent accuracy, so confirming whether that accuracy reflects genuine diagnostic learning is vital. This paper examines whether that accuracy reflects genuine diagnostic learning or is simply an artifact of how the datasets are built. The study compares six model families, namely Logistic Regression, Random Forest, Support Vector Machine, XGBoost, FT-Transformer, and TabNet, across three UCI AQ-10 age variants: Child (*n* = 290), Adolescent (*n* = 103), and Adult (*n* = 699). Most emphasis is put on separating the behavioral items that define the diagnostic label from the demographic covariates that do not, since the label itself is a threshold applied to those same items. The effectiveness of this separation is measured using a normalized Circularity Index alongside item-only and demographics-only accuracy. Evaluation of the results has shown that item-based accuracy stays at 93.2 to 100 percent even after the pre-computed *result* score column is removed, while demographics-only accuracy falls to 50.7 to 71.5 percent, at or below the majority-class baseline in every dataset, giving a mean Circularity Index of 0.947. The same pattern is confirmed on two independent domains outside autism screening: DASS-42 depression screening (*n* = 35,481, CI 0.984) and the Rosenberg Self-Esteem Scale (*n* = 39,076, CI 0.896). Lastly, a diagnostic heuristic named AutoCirc is introduced to flag this kind of circularity automatically using mutual information, reproducing the manual Circularity Index on three of five datasets, and a constructive eight-of-ten-item AQ-10 short form is proposed as a practical outcome of the same analysis. The results of the study confirm that high reported accuracy on these datasets reflects the scoring arithmetic of the screening instrument rather than a learned diagnostic pattern, though this does not call the AQ-10 into question as a clinical screening tool when used as intended. Further work shall seek to extend this audit to additional screening instruments, such as PHQ-9 and GAD-7, and to datasets carrying independent clinical diagnosis labels.

## I. Introduction

Autism Spectrum Disorder (ASD) affects an estimated 1 in 100 children worldwide, and early screening is central to timely intervention [1]. Machine learning has been applied to ASD detection using MRI [2], [3], EEG [4], and gait video [5]. Questionnaire-based screening, including the AQ-10 (child, adolescent, adult) and Q-CHAT-10 (toddler) instruments distributed through the UCI Machine Learning Repository, has attracted more than 150 published machine learning studies by an informal count [20]–[23]. Nearly all of these studies report 95 to 100 percent accuracy using Support Vector Machine (SVM), Random Forest, or XGBoost, yet none that we reviewed report demographics-only accuracy against the majority-class baseline, which is the comparison that exposes the circularity in this data. The high reported accuracy is not surprising once the dataset structure is examined. Each UCI AQ-10 record includes a *result* field, a weighted sum of the ten item scores, and the target label *Class: ASD* is simply a threshold applied to that same sum. A classifier given *result* needs only to learn this threshold rule rather than any genuine diagnostic pattern. Sollis et al. [18] confirmed this through a structured leakage audit named Clear-Rd, finding that the AQ-10 label is approximately determined by a score of 6 or higher, and that demographics-only performance falls to a Receiver Operating Characteristic Area Under the Curve (ROC-AUC) of 0.766 once the score is removed. What remains open is whether dropping *result* is enough to remove this problem. We show that it is not. The raw AQ-10 items are the components from which the label was computed, so any model with access to them can reconstruct the same rule without the score column. Practitioners who drop the result column may still believe their model has learned genuine diagnostic patterns, and papers comparing model families may in fact be measuring threshold-reconstruction fidelity rather than diagnostic capability. These problems are known conceptually [6], [19]. Our contribution is to quantify their severity, show that they persist after common mitigations, and provide multi-domain empirical evidence. This paper makes five contributions. First, a six-model, three-dataset demonstration that items-only accuracy of 93.2 to 100 percent stays at ceiling even after *result* is removed, extending the leakage audit of Sollis et al. [18] to the item level. Second, a normalized Circularity Index (mean AQ-10 value 0.947) that operationalizes the framework of Hagmann and Riezler [6] as a comparable scalar across datasets and models. Third, validation across three domains, adding DASS-42 depression screening (*n*=35,481, CI 0.984, with a comorbidity positive control) and the Rosenberg Self-Esteem Scale (*n*=39,076, CI 0.896). Fourth, AutoCirc, a mutual-information diagnostic heuristic, reported with its boundary-case and equal-weight failure modes disclosed rather than smoothed over. Fifth, an eight-of-ten-item AQ-10 short form retaining at least 95 percent of full accuracy, offered as a methodological consequence of circularity rather than a clinical recommendation.

### II. Related Work

It was found that Thabtah et al. [20] introduced a rule-based computational intelligence approach to autism screening, and this early work motivated the wide use of the AQ-10 and Q-CHAT-10 questionnaires as public machine learning benchmarks distributed through the UCI Machine Learning Repository. Rasul et al. [21] evaluated a range of classical classifiers on the same three age variants and reported accuracy of up to 100 percent, while Bala et al. [22] showed that ensemble methods such as Random Forest and boosting could reach 97.9 percent on the same data. Singh et al. [23] extended this with a four-axis benchmark adding deep tabular architectures (TabNet, FT-Transformer, TabPFN v2), finding near-ceiling accuracy was not limited to any single model family. Comparable accuracy has also been reported from EEG cepstral coefficients [16] and a broader mental-disorder diagnosis survey [17].

Despite this large body of work, almost none of these studies examine what the model is actually learning from the questionnaire. Sollis et al. [18] were among the first to look closely at this question, applying a structured leakage audit named Clear-Rd to the related Q-CHAT-10 toddler data and finding that the diagnostic label is close to a fixed threshold on the pre-computed score column, with demographics-only performance falling to a ROC-AUC of 0.766 once that column is removed. Hagmann and Riezler [6] had already described this general problem outside autism screening, defining circularity as a model’s apparent predictive power coming from reconstructing the arithmetic used to build its own label rather than from a genuine pattern in the data, and Kapoor and Narayanan [19] documented similar leakage across many machine-learning-based scientific fields, including NLP and tabular learning.

Table I makes the pattern easy to see: most studies report high accuracy but almost none test whether it survives once the items are excluded and only independent covariates remain, and the one audit that does, Sollis et al. [18], stops at the score column rather than the raw items it was built from. This is the gap addressed here: the same leakage check is extended from the score column to the item level, across three AQ-10 age variants and six model families, and repeated on two datasets outside autism screening.

**TABLE I.** Overview of related studies on AQ-10/Q-CHAT autism screening.

| Ref. | Methodology | Datasets Used | Outcome / Results | Limitations | Research Gap |
|---|---|---|---|---|---|
| [20] | Rule-based feature selection | AQ-10 Child, Adol., Adult (UCI) | Established AQ-10 as a public ML benchmark | Small samples; no clinical diagnosis label | Item-vs-label circularity not examined |
| [21] | Comparison of classical ML classifiers | AQ-10 Child, Adol., Adult | Up to 100% accuracy, all variants | No demographics-only baseline | Cannot separate signal from scoring rule |
| [22] | Ensemble learning (RF, boosting) | AQ-10 (UCI) | Up to 97.9% accuracy | Ensemble gain untested vs. item-only | Same circularity risk, untested |
| [23] | Four-axis benchmark: TabNet, FT-Transformer, TabPFN v2 | AQ-10 Child, Adol., Adult | High accuracy, classical & deep models | Questionnaire-derived, not clinical, labels | No item vs. demographic ablation |
| [18] | Clear-Rd leakage audit | Q-CHAT-10 toddler | Demo-only ROC-AUC 0.766 after score removed | Audit stops at the score column | Whether raw items reproduce the leakage |
| [6] | Formal circularity test | General, not screening-specific | Defines circularity as label-arithmetic reconstruction | Not applied to health-screening data | No domain-specific AQ-10/Q-CHAT use |

Our contribution is what we call structural circularity: a label defined as an arithmetic function of its own features, surviving removal of the score column and requiring domain knowledge to detect.

## III. Dataset Description

We use five publicly available datasets from two sources, summarized in Table II. The three AQ-10 age-variant datasets come from the UCI Machine Learning Repository under a CC BY 4.0 license [7]–[9] and share ten binary behavioral items. DASS-42 and the Rosenberg Self-Esteem Scale (RSE) are from the Open-Source Psychometrics Project [10], [12], verified against the published manual [11]. RSE items are scored 1–4, Strongly Disagree to Strongly Agree. Majority-class baselines in Table II are computed from the cleaned data used in our experiments.

**TABLE II.** Datasets used.

| **Dataset** | **Domain** | **N** | **Maj. base.** |
|---|---|---|---|
| AQ-10 Child | ASD, 4–11y | 290 | 51.7% |
| AQ-10 Adolescent | ASD, 12–16y | 103 | 60.2% |
| AQ-10 Adult | ASD, 17+ | 699 | 73.3% |
| DASS-42 Dep. | Depression, adult | 35,481 | 68.1% |
| RSE | Self-esteem, adult | 39,076 | 52.8% |

### A. Features in the Dataset

- **AQ-10 Behavioral Items**: ten binary (0/1) items shared across the Child, Adolescent, and Adult variants, each answered on the screening questionnaire and summed to produce the pre-computed *result* field that the label is thresholded from.
- **Demographic Covariates**: age, gender, ethnicity, country of residence, family history of autism, jaundice at birth, prior app use, and relation of the test-taker (parent, self, relative, health professional, or missing), none of which enter the item-scoring rule. Categorical covariates were one-hot encoded: ethnicity (11 categories) and relation (5 categories) without bucketing; country was collapsed into the five most frequent countries plus a single “Other” category.
- **DASS-42 Items**: 14 Depression items (rescaled 0–3) defining the label, plus 28 comorbid Anxiety and Stress items as a positive control, alongside age and gender. The Depression label is a score of at least 14, per the DASS manual [11].
- **RSE Items**: ten self-esteem items (scored 1–4, five reverse-coded) summed into an RSE total, alongside age and gender. The label defines low self-esteem as a total score of 25 or below, the standard midpoint of the instrument’s 10–40 scale [13].

For each dataset, instrument features are the items used to define the label, and covariate features are the demographic variables listed above, independent of the label’s construction. Continuous features were standardized inside each cross-validation fold to prevent leakage from scaling, and no feature selection or augmentation was applied. Missing or invalid responses were dropped at the record level, using the same procedure across all five datasets.

## IV. Proposed Methodology

With *A*_instr_, *A*_cov_, and *A*_base_ denoting the accuracy achieved using instrument features alone, covariate features alone, and the majority-class baseline, we define the Circularity Index in Equation 1.

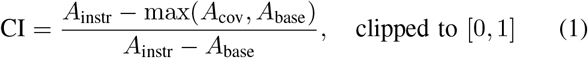

A Circularity Index approaching 1 indicates that predictive power sits entirely inside the instrument-defined features. A value approaching 0 indicates that covariates alone can match instrument accuracy, meaning genuine structure exists beyond the instrument’s own construction. We set CI to 0 whenever *A*_instr_ − *A*_base_ is 0.01 or smaller, to avoid instability from a near-zero denominator. CI is a diagnostic heuristic rather than a psychometrically validated measure. Unlike permutation importance, it summarizes the relative gap between feature groups rather than individual feature contributions. One consequence of the max(*A*_cov_, *A*_base_) term is that CI saturates at exactly 1.0 whenever *A*_cov_ ≤*A*_base_, regardless of how far below baseline covariates fall. This is why both AQ-10 Adult models report CI 1.000 in Table IV despite covariate accuracy sitting two points below baseline. The metric should be read alongside the raw *A*_cov_ and *A*_base_ values.

### A. Model Selection and Training

- **Logistic Regression**: linear baseline under nested cross-validation with a grid search over regularization strength.
- **Random Forest**: bagged decision-tree ensemble, testing nonlinear interactions.
- **SVM (RBF kernel)**: kernel-based nonlinear baseline alongside Random Forest.
- **XGBoost**: gradient-boosted tree ensemble, the primary nonlinear comparison to Logistic Regression.
- **FT-Transformer** [14]: compact tabular transformer (*d*_model_=16, two heads, two layers, dropout 0.2).
- **TabNet** [15]: attention-based tabular deep model (*n*_*d*_=*n*_*a*_=8, three decision steps).

For the four classical models, nested cross-validation was applied with 5 outer and 3 inner folds, running a grid search inside each outer fold so hyperparameter selection does not affect the test evaluation. The two deep tabular models used fixed architectures under plain 5-fold cross-validation, both trained from random initialization in each fold. Deep-model variance on the smaller AQ-10 datasets is high (fold-level SD up to 17 points), so these results are reported for completeness but the classical models under nested cross-validation are treated as the primary evidence. All experiments used a fixed random seed of 42, with Python 3.12, scikit-learn 1.5+, XGBoost 2.x, PyTorch 2.x+, and R 4.6.1.

#### Algorithm 1

AutoCirc: Automated Circularity Detection

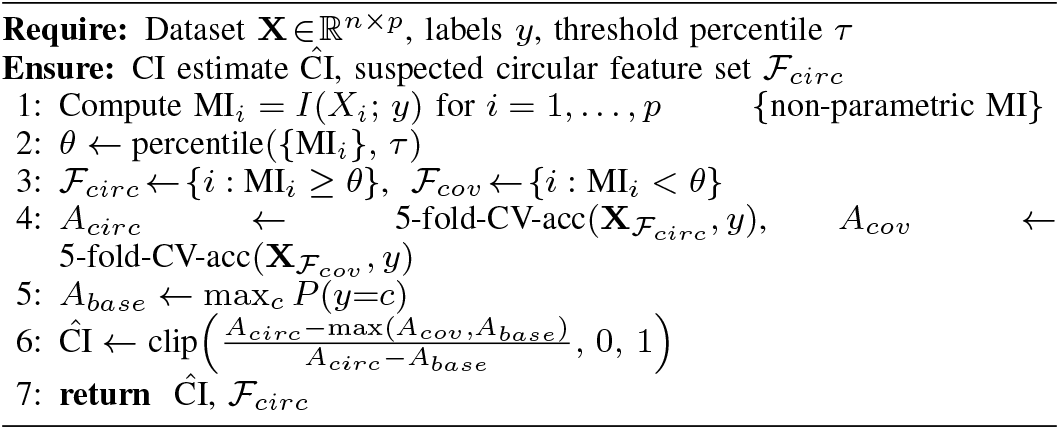

### B. Performance Metrics

- **Accuracy**: reported for instrument-only, covariate-only, and combined feature sets, since the gap between the first two exposes circularity.
- **Circularity Index**: the ratio in Equation 1, summarizing that gap as one comparable score across datasets and models.
- **Brier score**: checks calibration of the items-only and demographics-only models against the value expected from a random predictor.
- **SHAP importance**: corroborates the accuracy-based ablation and ranks items for the short-form result.
- **Item recall**: for AutoCirc, the fraction of true instrument items correctly flagged as circular.

AutoCirc, described in Algorithm 1, is a zero-knowledge screening heuristic that uses mutual information as a proxy for label alignment, flagging potentially circular features before a full ablation study is run. At the default threshold percentile of 50 (a median split), it requires no domain knowledge and completes in under 60 seconds on the largest dataset here, RSE (*n*=39,076).

## V. Results

### A. Single-Dataset Ablation (AQ-10 Child)

Table III reports accuracy for all six model families on AQ-10 Child across five feature-set conditions. Across the four classical models and FT-Transformer, both the items-only and items-plus-demographics conditions stay within 93.4 to 100.0 percent even after the *result* field is removed, with Logistic Regression reaching 99.3 percent on items only (SD 1.9 points). For LR, SVM, and XGBoost, adding demographics to the items slightly reduces accuracy rather than improving it (e.g., LR falls from 99.3 to 97.6 percent). With only *n*=290 records, the added demographic features contribute noise rather than signal once the items already carry the label’s arithmetic. TabNet is more variable (85.9 percent items-only, 72.4 percent items-plus-demographics), consistent with the higher fold-level variance noted for deep models in Section IV. Once AQ-10 items are excluded entirely, accuracy collapses to 50.7–55.9 percent across all six models, at or below the 51.7 percent majority-class baseline (LR: 55.9 percent, SD 8.1 points). Fig. 1 shows this pattern for the best model per condition: accuracy stays near ceiling whenever AQ-10 items are present and drops sharply once they are removed.

**TABLE III.** AQ-10 Child accuracy by feature set and model (LR–XGB: nested CV; FT-Tr./TabNet: fixed arch.).

| **Feat. set** | **LR** | **RF** | **SVM** | **XGB** | **FT-Tr.** | **TabNet** |
|---|---|---|---|---|---|---|
| result only | 100.0 | 100.0 | 100.0 | 100.0 | – | – |
| items+demo+result | 100.0 | 100.0 | 100.0 | 100.0 | – | – |
| items only | 99.3 | 93.4 | 98.3 | 97.2 | 100.0 | 85.9 |
| items+demo | 97.6 | 93.4 | 95.2 | 96.9 | 94.8 | 72.4 |
| demo only | 55.9 | 52.8 | 54.8 | 54.8 | 52.4 | 50.7 |

**Fig. 1.**
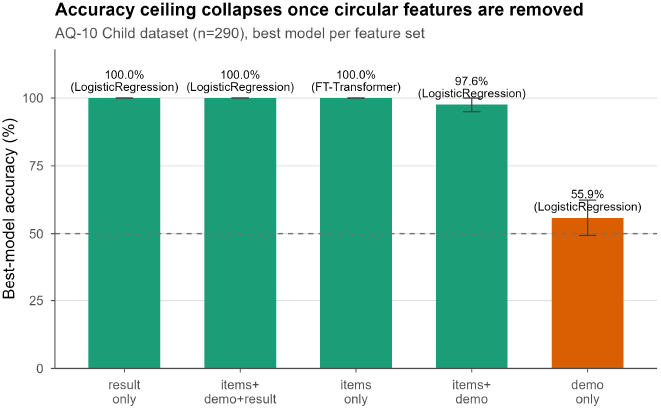
Accuracy by feature set, AQ-10 Child.

### B. Cross-Dataset Generalization and Circularity Index

This pattern holds consistently across all three AQ-10 age variants, as shown in Fig. 2, which plots Logistic Regression accuracy across the three datasets. Item-containing feature sets are shown in green, demographics-only accuracy in orange, and each dataset’s majority baseline as a dashed line. In the Adult dataset the result is especially notable: demographics-only models score below the 73.3 percent majority baseline, at 71.5 percent for Logistic Regression and 70.5 percent for XGBoost, confirming that demographic features carry no genuine signal once class imbalance is taken into account. Table IV summarizes the Circularity Index, computed using Equation 1, across all five datasets. The mean Circularity Index across the six AQ-10 rows in this table is 0.947.

**TABLE IV.**
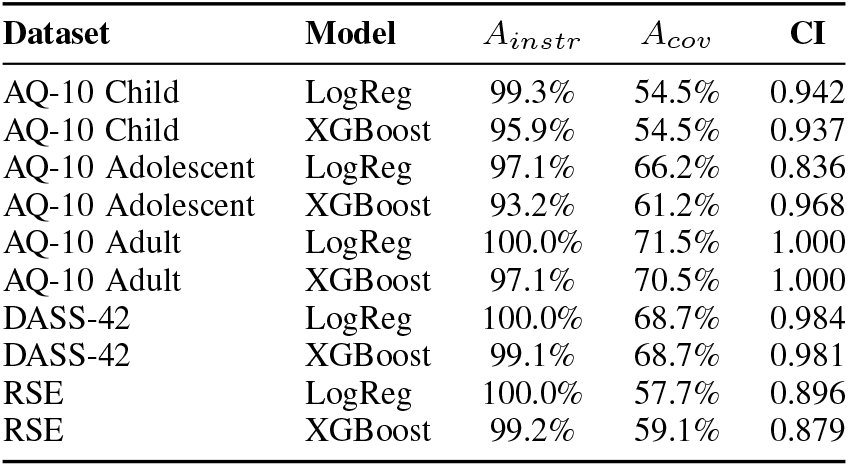
Circularity Index Across Datasets.

| **Dataset** | **Model** | *A*<sub>*instr*</sub> | *A*<sub>*cov*</sub> | **CI** |
|---|---|---|---|---|
| AQ-10 Child | LogReg | 99.3% | 54.5% | 0.942 |
| AQ-10 Child | XGBoost | 95.9% | 54.5% | 0.937 |
| AQ-10 Adolescent | LogReg | 97.1% | 66.2% | 0.836 |
| AQ-10 Adolescent | XGBoost | 93.2% | 61.2% | 0.968 |
| AQ-10 Adult | LogReg | 100.0% | 71.5% | 1.000 |
| AQ-10 Adult | XGBoost | 97.1% | 70.5% | 1.000 |
| DASS-42 | LogReg | 100.0% | 68.7% | 0.984 |
| DASS-42 | XGBoost | 99.1% | 68.7% | 0.981 |
| RSE | LogReg | 100.0% | 57.7% | 0.896 |
| RSE | XGBoost | 99.2% | 59.1% | 0.879 |

**Fig. 2.**
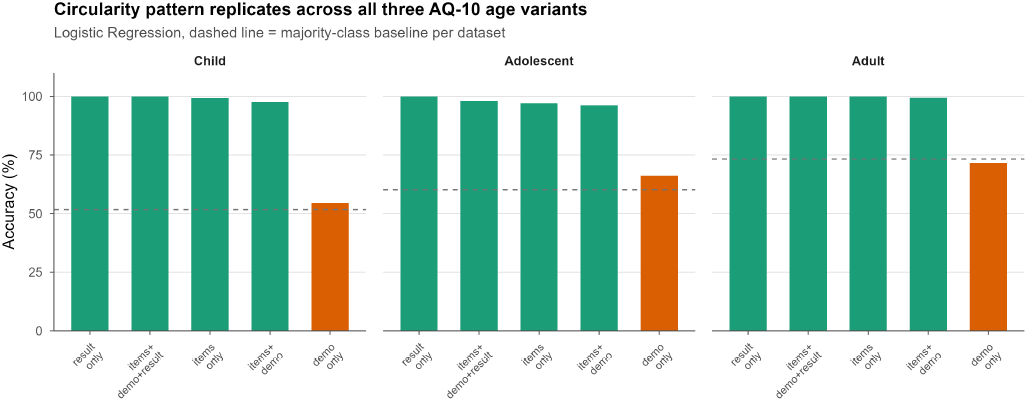
Accuracy across AQ-10 age variants (Logistic Regression).

### C. Cross-Domain Validation (DASS-42 and RSE)

On the cleaned DASS-42 dataset (*n*=35,481), Depression items alone reach 100.0 and 99.1 percent accuracy for Logistic Regression and XGBoost against a 68.1 percent majority baseline (CI 0.984 and 0.981). The 28 comorbid Anxiety and Stress items, not part of the Depression label’s definition, reach 83.0 and 83.5 percent, 14.9 points above baseline. This positive control rules out modeling failure as the explanation for the near-baseline demographics collapse, shown in the left panel of Fig. 3. On RSE (*n*=39,076), the ten self-esteem items reach 100.0 and 99.2 percent against a 52.8 percent baseline (CI 0.896 and 0.879), while demographics (age, gender) reach only 57.7 and 59.1 percent, shown in the right panel of Fig. 3. Repeating this at RSE thresholds of 15, 20, 25, and 30 gives a Circularity Index ranging from 0.886 to 1.000. Together with the AQ-10 findings, these two domains establish a three-domain empirical pattern: instrument-defining items give 93 to 100 percent accuracy, while independent demographics collapse to the majority baseline.

**Fig. 3.**
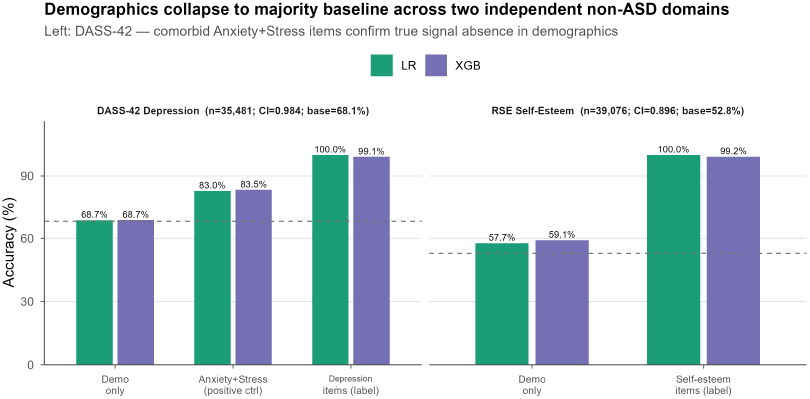
Cross-domain validation, DASS-42 and RSE.

### D. AutoCirc Validation

Table V reports AutoCirc, run at the median-split threshold, on all five datasets. On AQ-10 Adolescent, AQ-10 Adult, and DASS-42, the estimated Circularity Index is 0.987 or higher, with item recall between 80 and 100 percent. On AQ-10 Child, a boundary-case failure occurs because the A2 Score item sits at the edge of the mutual-information cluster, lowering the estimated index to 0.664 despite a manual Circularity Index of 0.942, shown in Fig. 4 (AQ-10 items in teal, demographics in orange). Items cluster above a mutual-information value of about 0.04, and demographics fall below it, except for this boundary case. RSE reveals a second failure mode. Because all ten items contribute equally to the total, they carry similar mutual information, and the median-split threshold assigns only 7 of 10 to the circular feature set (70 percent recall). The remaining three, though still circular, raise covariate accuracy to 83.5 percent and deflate the estimated index to 0.286 against a manual CI of 0.896. These two failure modes show that AutoCirc performs best when circular and non-circular features form a clear mutual-information gap. This gap is met by AQ-10 Adult, Adolescent, and DASS-42, but not equal-weight instruments such as RSE. On two datasets with no known circularity, the Wisconsin Breast Cancer set (*n*=569) correctly returns a low estimated CI of 0.517 (covariates remain 15.3 points above baseline), while Adult Census Income (*n*=48,842) gives a false-positive estimate of 0.967, caused by class imbalance shrinking the denominator in Equation 1 to 5.6 points. This confirms that domain knowledge remains necessary to distinguish genuine circularity from imbalance artifacts.

**Table V.** AutoCirc versus manual Circularity Index.

| **Dataset** | **N** | $\hat{A}_{circ}$ | $\hat{A}_{cov}$ | **Auto CI** | **Manual CI** | **Item rec.** |
|---|---|---|---|---|---|---|
| AQ-10 Child | 290 | 93.8% | 65.9% | 0.664 | 0.942 | 90% |
| AQ-10 Adolescent | 103 | 93.2% | 60.2% | 0.999 | 0.836–0.968 | 80% |
| AQ-10 Adult | 699 | 95.9% | 73.5% | 0.987 | 1.000 | 90% |
| DASS-42 | 35,481 | 100.0% | 68.5% | 0.988 | 0.981–0.984 | 100% |
| RSE | 39,076 | 95.7% | 83.5% | 0.286 | 0.896 | 70% |

**Fig. 4.**
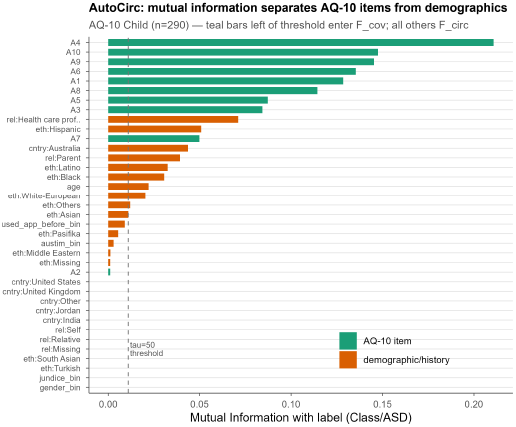
AutoCirc mutual information profile, AQ-10 Child.

### E. Interpretability, Constructive Shortening, and Fairness

SHAP analysis on the AQ-10 Child model with *result* removed assigns 95.8 percent of total feature importance to the AQ-10 items and only 4.2 percent to demographics (Fig. 5), independently corroborating the ablation results. Per-sample 5-fold cross-validation on AQ-10 Child yields only 2 misclassifications out of 290 (0.69 percent), both at the exact label threshold of a score of 6, confirming the scoring rule is fully recoverable except at this single ambiguous boundary. Brier scores agree. The items-only Logistic Regression model scores 0.013 (near-perfect calibration) versus 0.262 for demographics-only, close to the 0.250 expected from a random predictor. Training on the top *k* SHAP-ranked items for *k* = 1 to 10, the smallest subset retaining at least 95 percent of full accuracy is *k* = 8 (95.7 percent), dropping only A3 and A6. This is a methodological consequence of the instrument’s own scoring arithmetic rather than a clinical recommendation. A gender subgroup fairness audit on the result-removed model (Fig. 6) finds 97.6 percent accuracy for female subjects (*n*=84) versus 93.7 percent for male subjects (*n*=206), 94.8 percent overall. Given the 71:29 gender imbalance in this dataset, this gap should be monitored rather than assumed away, since it reflects differences in item-response patterns between genders rather than differential diagnostic capability. This distinction matters for resource-allocation decisions.

**Fig. 5.**
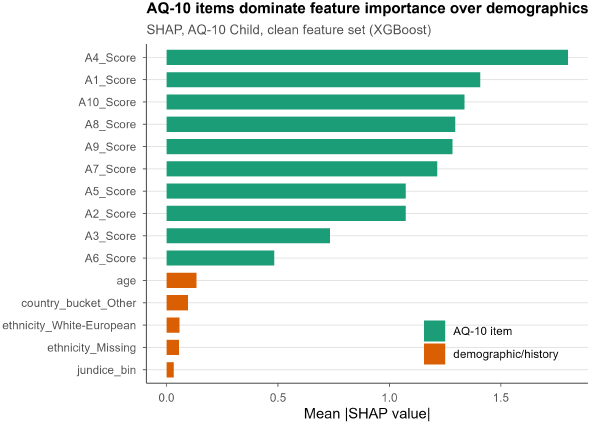
SHAP feature importance, AQ-10 Child.

**Fig. 6.**
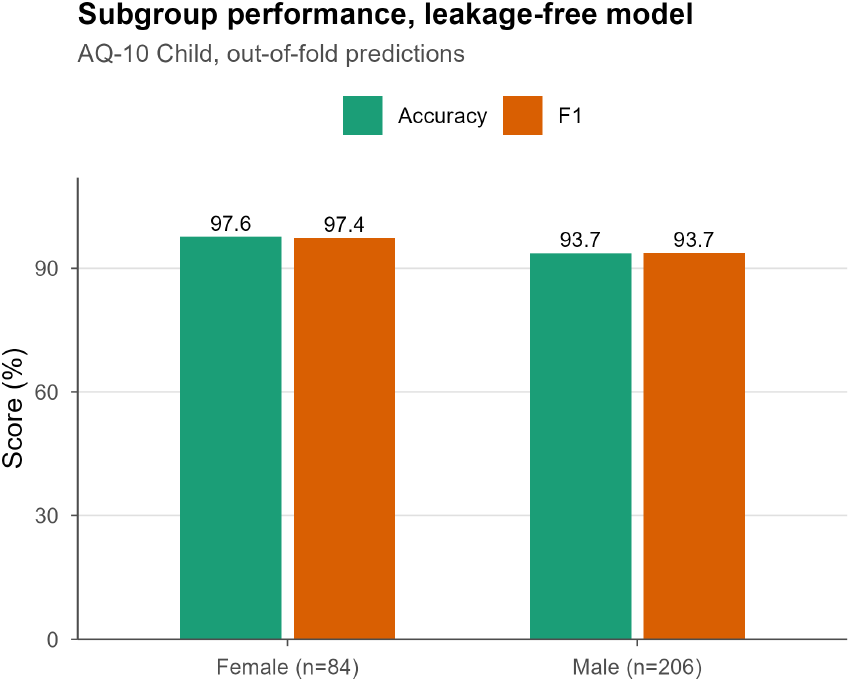
Subgroup accuracy by gender.

## VI. Discussion

### A. Interpretation and Practical Implications

Prior claims of 95 to 99 percent accuracy on AQ-10 and Q-CHAT datasets reflect reconstructed scoring arithmetic, not a generalizable diagnostic pattern, even after *result* is removed. Researchers building genuine ASD predictive models should use independent clinical diagnosis labels, such as ADOS-2, so the labeling process stays independent of the scored instrument. Any study on questionnaire data should verify the label is not an arithmetic function of the predictors, report demographics-only accuracy against baseline, and run AutoCirc as a zero-knowledge pre-study audit. Beyond the boundary-case and equal-weight failure modes already described in Section V-D, three further constraints apply: correlated predictors can share mutual information in ways that mask individual circularity, weighted or nonlinear scoring rules cause mutual information to reflect distributional weight rather than structural circularity, and severe class imbalance shrinks the denominator in Equation 1, pushing the Circularity Index toward 1 even without genuine circularity, as observed for Adult Census Income. Domain knowledge thus remains essential for confirming any result AutoCirc produces.

### B. Limitations

Internal validity is limited by the small size of the Child (*n*=290) and Adolescent (*n*=103) datasets, so deep-model results on them should be read as exploratory. The circularity finding itself is model-agnostic and confirmed by the classical models under full nested cross-validation, and fixed hyperparameters for the deep models reflect available compute. All experiments used a single fixed seed (42) with no multi-seed repeats, so reported accuracy and SD values do not reflect seed-to-seed variance. Sparse one-hot columns on the smaller datasets may also constrain the demographics-only model’s ability to detect genuine covariate signal, so the demographics collapse is best read as evidence against signal rather than proof of its total absence. Per-fold SD is reported precisely only for Logistic Regression; for FT-Transformer and TabNet we report only the ceiling value (17 points), an asymmetry we flag rather than resolve here. External validity is limited because the 150-plus published-study count comes from an informal web and PubMed search, the DASS-42 and RSE samples are self-selected online respondents who may not represent clinical populations, and the Q-CHAT-10 toddler variant on UCI was not accessible. Construct validity is limited because all labels reflect screening-tool outcomes rather than independent clinical diagnoses, and CI saturates at 1.0 whenever covariates fall at or below baseline, so it should be read alongside its raw accuracy inputs. The RSE threshold of 25 is a standard midpoint cutoff; our sensitivity analysis in Section V-C confirms stability across cutoffs from 15 to 30 (CI 0.886–1.000). AutoCirc’s median-split threshold (*τ* = 50) was not swept, so whether the RSE equal-weight failure mode is threshold-dependent remains untested.

## VII. Conclusion

Across six model families and three UCI AQ-10 age variants, we have shown that dropping the pre-computed *result* score does not resolve label circularity, because the behavioral items themselves define the label as a near-deterministic threshold function, with a mean Circularity Index of 0.947, extending the leakage audit of Sollis et al. [18] to the item level. Two independent domains, DASS-42 depression screening (CI 0.984, with a built-in comorbidity positive control) and the Rosenberg Self-Esteem Scale (CI 0.896), confirm the same pattern across ASD screening, clinical depression, and general psychometric self-report. Researchers building genuine ASD predictive models should use independent clinical diagnosis labels, such as ADOS-2, as the prediction target, keeping the labeling process separate from the scored instrument. Until that separation is standard practice, accuracy figures in this space measure threshold-reconstruction fidelity rather than diagnostic capability. We offer AutoCirc as a lightweight pre-study check and encourage its application to further instruments, including PHQ-9 and GAD-7.

## Data Availability

All data used in this study are publicly available from the UCI Machine Learning Repository: Autistic Spectrum Disorder Screening Data for Children, for Adolescents, and for Adults (CC BY 4.0).

https://archive.ics.uci.edu/dataset/419

https://archive.ics.uci.edu/dataset/420

https://archive.ics.uci.edu/dataset/426

## References

[1] World Health Organization, “Autism spectrum disorders” fact sheet, 2023. [Online]. Available: https://www.who.int/news-room/fact-sheets/detail/autism-spectrum-disorders

[2] H. S. Nogay and H. Adeli, “Multiple classification of brain MRI autism spectrum disorder by age and gender using deep learning,” J. Med. Syst., vol. 48, no. 1, p. 15, 2024. doi: 10.1007/s10916-023-02032-0

[3] V. R. P. K., C. H. Bindu, and K. R. Devi, “An interpretable deep learning approach for autism spectrum disorder detection in children using NASNet-mobile,” Biomed. Phys. Eng. Express, vol. 11, no. 4, art. 045006, 2025. doi: 10.1088/2057-1976/addbe7

[4] F. Abedinzadeh Torghabeh, Y. Modaresnia, and M. H. Moattar, “Hybrid deep transfer learning-based early diagnosis of autism spectrum disorder using scalogram representation of electroencephalography signals,” Med. Biol. Eng. Comput., vol. 62, no. 2, pp. 495–503, 2024. doi: 10.1007/s11517-023-02959-z

[5] F. Colonnese, F. Di Luzio, A. Rosato, and M. Panella, “Bimodal feature analysis with deep learning for autism spectrum disorder detection,” Int. J. Neural Syst., vol. 34, no. 2, 2450005, 2024. doi: 10.1142/S0129065724500059

[6] M. Hagmann and S. Riezler, “False perfection in machine prediction: Detecting and assessing circularity problems in machine learning,” 2021. 2106.12417

[7] UCI Machine Learning Repository, “Autistic Spectrum Disorder Screening Data for Children,” 2017, CC BY 4.0. [Online]. Available: https://archive.ics.uci.edu/dataset/419

[8] UCI Machine Learning Repository, “Autistic Spectrum Disorder Screening Data for Adolescent,” 2017, CC BY 4.0. [Online]. Available: https://archive.ics.uci.edu/dataset/420

[9] UCI Machine Learning Repository, “Autism Screening Adult,” 2017, CC BY 4.0. [Online]. Available: https://archive.ics.uci.edu/dataset/426

[10] Open-Source Psychometrics Project, “DASS raw data, 2017–2019.” [Online]. Available: https://openpsychometrics.org/_rawdata/DASS_data_21.02.19.zip

[11] S. H. Lovibond and P. F. Lovibond, Manual for the Depression Anxiety Stress Scales, 2nd ed. Sydney: Psychology Foundation, 1995.

[12] Open-Source Psychometrics Project, “Rosenberg Self-Esteem Scale (RSE) raw data.” [Online]. Available: https://openpsychometrics.org/_rawdata/RSE.zip

[13] M. Rosenberg, Society and the Adolescent Self-Image. Princeton, NJ: Princeton University Press, 1965.

[14] Y. Gorishniy, I. Rubachev, V. Khrulkov, and A. Babenko, “Revisiting deep learning models for tabular data,” in Proc. NeurIPS, 2021. 2106.11959

[15] S. O. Arik and T. Pfister, “TabNet: Attentive interpretable tabular learning,” in Proc. AAAI, 2021. doi: 10.1609/aaai.v35i8.16826

[16] R. Menaka, R. Karthik, S. Saranya, M. Niranjan, and S. Kabilan, “An improved AlexNet model and cepstral coefficient-based classification of autism using EEG,” Clin. EEG Neurosci., vol. 55, no. 1, pp. 43–51, 2024. doi: 10.1177/15500594231178274

[17] F. D. E. Baran and M. Cetin, “AI-driven early diagnosis of specific mental disorders: a comprehensive study,” Cogn. Neurodyn., vol. 19, no. 1, p. 70, 2025. doi: 10.1007/s11571-025-10253-x

[18] L. J. Sollis, D. P. Wall, and P. Y. Washington, “Evaluating multicultural autism screening for toddlers using machine learning on the QCHAT-10,” medRxiv, Nov. 2024. doi: 10.1101/2024.11.12.24317211

[19] S. Kapoor and A. Narayanan, “Leakage and the reproducibility crisis in machine-learning-based science,” Patterns, vol. 4, no. 9, 100804, 2023. doi: 10.1016/j.patter.2023.100804

[20] F. Thabtah, F. Kamalov, and K. Rajab, “A new computational intelligence approach to detect autistic features for autism screening,” Int. J. Med. Inform., vol. 117, pp. 112–124, 2018. doi: 10.1016/j.ijmedinf.2018.06.009

[21] R. A. Rasul, P. Saha, D. Bala, S. M. R. U. Karim, M. I. Abdullah, and B. Saha, “An evaluation of machine learning approaches for early diagnosis of autism spectrum disorder,” 2023. 2309.11646

[22] M. Bala, M. H. Ali, M. S. Satu, K. F. Hasan, and M. A. Moni, “Efficient machine learning models for early stage detection of autism spectrum disorder,” Algorithms, vol. 15, no. 5, p. 166, 2022. doi: 10.3390/a15050166

[23] S. Singh et al., “ASD-Bench: A four-axis comprehensive benchmark of AI models for autism spectrum disorder,” 2026. 2605.11091

